# The impact of multiplex panel testing on ascertainment of pertussis-attributable deaths: national surveillance from England

**DOI:** 10.64898/2025.12.03.25341094

**Authors:** Catherine Sandford Alber, Emer Cullen, Sonia Ribeiro, David J. Litt, Gayatri Amirthalingam, Helen Campbell, Sharif A. Ismail

**Author notes:** **CONTACT:** Dr. Sharif A. Ismail, Consultant Epidemiologist, UK Health Security Agency, 61 Colindale Avenue, London NW9 5EQ, E.

## Abstract

Understanding the burden of pertussis and pertussis-related mortality is important in enabling better understanding of population vulnerability and supporting vaccination programme planning. However, changes in laboratory practice can have important implications for public health surveillance. This study considers the impact of increasing use of multiplex panel testing for respiratory pathogens on ascertainment of pertussis-related deaths, drawing on multiple surveillance data sources from England. Information was sourced from Office of National Statistics cause of death records, public health case management records, laboratory results collated nationally, and death notifications received via General Practices. Figures covered the period 2023/24, coinciding with a large, national outbreak in England, and were compared with data from 2012 – the last major outbreak in England. Of 34 deaths identified among individuals aged >12 months in 2023/24, 11 ultimately met pre-defined criteria to be classified as pertussis-related deaths; none of the 5 possible cases identified in 2012 did so. A total of 25 of the 34 (74%) tested positive for pertussis via multiplex panel testing, of which 18 (72%) were ultimately excluded as pertussis-related deaths. Findings highlight significant challenges in attribution of pertussis-related mortality in an era of widespread multiplex panel testing.

## Introduction

Pertussis is endemic worldwide and despite the implementation of long-standing vaccination programmes, it remains a cyclical disease with peaks in cases occurring every three to five years in England (1). The primary objective of the pertussis immunisation programme in England is to prevent deaths in infants, the highest risk group for severe outcomes. There is more limited data internationally on the risk of pertussis in older children, adolescents and adults, and reliably assessing the true morbidity and mortality attributable to pertussis is more challenging.

Measures introduced during the COVID-19 pandemic resulted in very low rates of pertussis in 2021 and 2022 in England (2) and indeed across Europe (3). In England, there was a surge in cases across all age groups from summer 2023 (4) continuing into 2024, with 14,894 laboratory confirmed cases in 2024 (5). This was the largest outbreak in England since 2012, which at the time had been the largest increase in pertussis cases in over 20 years (6). The 2023/24 outbreak was also the largest in England since the introduction of the maternal vaccination programme, commenced in October 2012 (6). A similar increase was seen across Europe, with notification rates recorded across the European Union in 2023 ranging from 0 to 77.53 per 100,000 (7).

Routine surveillance for pertussis cases is carried out across Europe to monitor circulating pertussis levels, and several European countries routinely report on pertussis-related deaths. The European Centre for Disease Prevention and Control annual pertussis report for 2023 documents 10 deaths across the continent affecting infants and those in older age groups (7). In England, infant deaths from pertussis are routinely reported and data on these deaths form an important part of ongoing work to monitor the effectiveness of maternal and infant pertussis vaccination programmes (4).

The process for attribution of deaths due to pertussis is complex. In England, a key source is death certification from the Office for National Statistics (ONS), but in some cases final judgement on cause of death may be delayed (e.g. where a post-mortem or case investigation may be required). Laboratory testing is central, but there have been important changes in many countries in testing approaches prior to, and since the pandemic which influence mortality attribution.

The most common laboratory techniques for detecting *Bordetella pertussis* infections are culture, polymerase chain reaction (PCR), and immunological assays (e.g. enzyme-linked immunosorbent assay (ELISA) and multiplexed immunoassays (MIA)) for serological testing (8, 9). Testing approaches vary by age group – PCR being the mainstay for case confirmation in infants and younger age groups. PCR offers a rapid test result with high sensitivity, although there is a small chance of false positives with some gene targets (8). However, multiplex PCR panels are now widely used in England, and in many other high income countries worldwide (10). Multiplex panel testing involves the simultaneous testing of a single sample for multiple pathogens using PCR. *B. pertussis* PCR is included in a range of respiratory multiplex PCR panels alongside common respiratory viruses such as influenza, RSV and SARS-CoV-2. The use of multiplex panels can reduce the sensitivity for pertussis detection (11, 12) but may also result in detection of cases that would not have otherwise been tested for pertussis due to atypical presentation or lack of clinical identification, the latter being more likely to occur in non-infant age groups.

The analysis reported in this paper aimed to assess the impact of the increasing use of multiplex panel testing on pertussis-related death ascertainment. We analysed data on non-infant pertussis deaths in England from the 2023/24 outbreak and compared findings with data from the last major outbreak in 2012 – a period when multiplex panel testing was not available.

## Methods

### Data sources

The UK Health Security Agency (UKHSA) collects routine surveillance on any deaths which may be related to *B. pertussis* infection. Non-infant pertussis related deaths are collated from the following sources:

- Notified cases (regardless of laboratory status) and cases with laboratory confirmation are collated by regional health protection teams and recorded on the national Case and incident management system (CIMS) on an individual level (formerly HPZone until summer 2024, then transitioned to CIMS). These are reviewed for information indicating a death;
- Deaths with pertussis as an underlying cause recorded by the Office for National Statistics (ONS) in cause of death (COD) records;
- Laboratory confirmed cases of pertussis where a death is recorded in the NHS Personal Demographic Service in temporal association with the laboratory result. These laboratory results may be from the national reference laboratory at Colindale, UKHSA regional laboratories via Data Mart or from local NHS laboratories with results recorded centrally on the second-generation surveillance system (SGSS), the national laboratory reporting system used in England to collect data on infectious diseases;
- Death notification via the enhanced surveillance form that is disseminated to the General Practitioners (GPs) that each laboratory confirmed case is registered with.

Where cause of death was not reported in ONS or was not clear, further information on cause of death was obtained from the National Childhood Mortality Dataset (NCMD) for cases aged under 18 years.

The collated lists of possible pertussis deaths for 2012 and from January 2023 to December 2024 were reviewed to compare cases who had died from the two most recent outbreaks. The CIMS/HPZone record was reviewed for each case to gather additional information. In cases where there was insufficient information on cause of death or test results were unavailable, the registered GP and, if required, the trust-based microbiologist or local coroner’s office, were contacted for further information

### Outcome definitions

Pertussis-related deaths were defined in this analysis as those in individuals aged over 12 months of age with either:

1. Primary definition: pertussis listed in the definitive COD from the ONS as part of the illness episode leading to death; or, where this information was not available:
2. Secondary definitions: EITHER (i) a positive pertussis laboratory test result with clinically compatible symptoms OR, in the absence of a test result, (iia) clinically compatible symptoms and no more likely diagnosis or (iib) clinically compatible symptoms and equivalently possible diagnosis but also a pertussis epidemiological link per case definition.

### Further investigation and analysis

For cases occurring in 2023/24 with a positive pertussis test result, the relevant hospital laboratory was contacted to determine the type of testing used and the clinical indication for pertussis testing. If applicable, laboratories were asked for the type of multiplex panel testing used and checked for UKAS accreditation. Cases with a positive test result from 2012 were assumed to not have used multiplex panel testing, as they occurred prior to the introduction of multiplex panel testing in hospital laboratories. Data from all sources above were collated and analysed descriptively in MS Excel.

## Results

In 2012, there were five non-infant deaths identified through routine surveillance routes as possible *B. pertussis* related deaths. The deaths were detected through laboratory reporting and GP notifications. The median age was 69 years (all deaths were in individuals in the age range between 55-59 and 70-74 years). None of the deaths met the criteria to be defined as a pertussis related deaths on review of ONS cause of death and the clinical information from HPZone notes and GP records (i.e. they did not meet the requirement for clinically compatible symptoms to be present). The laboratory confirmation method for all the cases was via serology.

In 2023/24, there were 34 non-infant deaths identified as possible pertussis related deaths. Nineteen were reported through pertussis database laboratory results, one from GP, seven from ONS death records and seven from CIMS/HPZone (**Figure 1**). The median age was 75 years (all deaths were in individuals in the age range between 5-9 and 90-94 years). From the deaths identified, 30 (88%) had an ONS cause of death available. Of these, 10 (29%) met the primary definition of pertussis being listed as a contributory cause of death, of which two deaths in adults were excluded because primary infection occurred many years previously, in childhood. For the four cases where ONS cause of death was unavailable, two did not meet the case definition that pertussis was a cause of death from the additional information acquired. For the remaining cases, one had cause of death still to be determined (and was therefore excluded), and one was confirmed to be a pertussis related death by the coroner following postmortem examination, giving a total of 11 pertussis related deaths in 2023/24. For the 11 deaths for which pertussis was considered a likely underlying cause, the median age was 75 years (all deaths were in individuals in the age range between 30-34 and 80-84 years). None of the cases had pertussis listed as the only cause of death. Most cases had at least one other chronic condition listed, with chronic respiratory illness being listed for four out of the 11 cases. Six of the cases had an additional respiratory pathogen listed.

**Figure 1:**
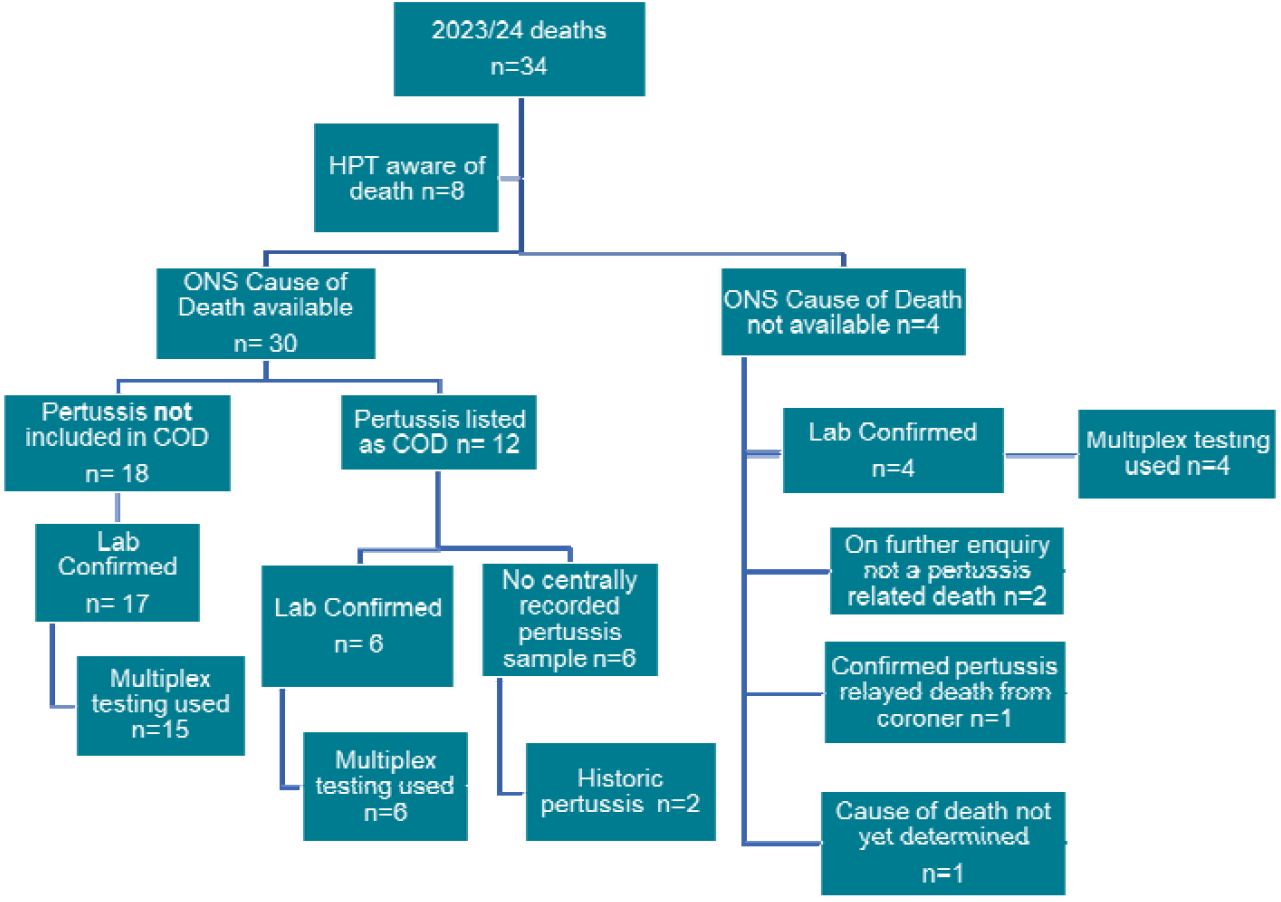
Flow-chart for notified possible pertussis-related deaths in England 2023/24 with ensuring breakdown outlining numbers laboratory-confirmed, and those testing positive specifically via multiplex panel testing. [Abbreviations: COD = Cause of Death; HPT = Health Protection Team; ONS = Office of National Statistics]

In 2023/24, of the 26 cases with a confirmed positive pertussis result centrally recorded, 23 (88%) had been tested using a multiplex panel. Out of these 23 cases, 16 did not meet the primary or secondary definition of a pertussis related death. In total 13 hospital microbiology laboratories were contacted (covering all those laboratories from which one or more positive pertussis results were recorded), seven of which had UKAS accreditation for multiplex panel testing. Some of the laboratories received samples from other hospitals and acted as the central site offering multiplex panel testing. A range of platforms were used including Qiagen Qiastat, Biomerieux Biofire and Aus Diagnostics. The criteria for use of multiplex panel testing varied across the country and included immunocompromise, high dependency- or intensive care unit admission, paediatric admission and severe illness. Many laboratories required microbiology consultant approval to use multiplex panel testing, but one laboratory routinely tested all throat swabs.

In those with pertussis listed as a COD by the ONS, six had no central record of a positive pertussis test result. On further investigation, four of the cases had records of a positive pertussis result in the local laboratory but this was not recorded centrally on SGSS. The remaining two cases did not have a positive pertussis result close to their date of death but had chronic complications from historic pertussis infection in childhood which were recorded as having contributed to their cause of death as adults.

## Discussion

Understanding the true incidence of pertussis and pertussis-related mortality can help to highlight areas of unmet need or vulnerable populations, and support vaccination planning (13). Changes in detection and case management approaches between the 2012 and the 2023/2024 outbreak mean that comparison of the number of adult deaths in the two time periods should be undertaken with caution. In the 2012 outbreak in England, we identified no deaths meeting the pertussis-related death definition applied in this analysis, compared to 11 deaths in 2023/24 that did meet the definition. Importantly, of 34 deaths potentially attributable to pertussis in 2023/24, 25 (74%) tested positive via multiplex panel testing, of which 18 (72%) were ultimately excluded as pertussis-related deaths according to the primary or secondary definitions used in this analysis.

Nearly half (45%) of the pertussis related deaths in England in 2023/24 were in individuals aged over 80 years of age and the majority were aged over 60 years. All cases had more than one cause of death listed in addition to pertussis. Most cases had another chronic illness, with chronic respiratory illness being the most common additional cause of death. However, given small numbers and the fact that these deaths occurred during a period of very high transmission in England with exceptionally high case numbers reported overall, these numbers should be interpreted with caution.

In particular, findings from this analysis underscore challenges in attribution of pertussis-related mortality in an era of widespread, extended multiplex panel testing, including for pertussis. Multiplex panel testing is now widely used in hospital laboratories in England. While international data are sparse, studies suggest up to a third of paediatric patients presenting with respiratory symptoms are tested in this way in some settings (14, 15). Studies have also shown that some multiplex PCRs show only 56-67% positive pertussis rates compared to those identified by single pertussis PCR testing in a cohort of clinically confirmed pertussis patients (8, 11, 12, 16, 17).

The use of multiplex panel testing increases the number of pertussis tests carried out, and this may have led to an increase in case detection and reporting in the 2023/24 outbreak (10, 18). In addition to the use of multiplex panel testing, active case finding and higher clinical awareness in the 2023/4 outbreak may also have contributed to the higher rates of pertussis-related death detection. It is also notable that the criteria used by many of the hospitals for multiplex panel testing (see above) may make the detection of pertussis more likely in individuals with other medical conditions or co-morbidities.

While 55% of the pertussis-related deaths also had another respiratory pathogen listed as a cause of death, it is difficult to determine from this analysis how far each individual infection (or co-infection with multiple pathogens as a distinct risk factor in itself) contributed to death in each case. The use of respiratory multiplex panel testing could increase the likelihood of incidental positive results. And when multiplex panel testing identifies more than one positive organism, it may be difficult to determine whether the infections identified co-contributed to the death or were an incidental finding, depending on the clinical presentation.

Our analysis also points to broader challenges in deaths surveillance. Obtaining accurate clinical information from GPs was sometimes challenging, particularly for cases where there was uncertainty over the cause of death. Several GPs mentioned that they do not receive any information from the coroners regarding the outcome of cause of death decisions. In addition, several deaths listed by the ONS with pertussis as a cause of death were found to have no central positive pertussis test result recorded in SGSS. On further investigation, the majority (excluding two historic pertussis cases) did have a positive pertussis result locally, highlighting a potential issue in the transfer of data from local laboratories to SGSS. This may in part be due to increased use of PCR testing, with fewer cultures being sent to the reference laboratory. It requires further investigation to ensure that notifications, and potential public health actions, are not being missed.

This analysis highlights challenges in attribution of pertussis-related mortality in an era of widespread multiplex panel testing. The most recent pertussis outbreak (2023/24) saw a higher number of pertussis related deaths than in 2012. This may be the result of changes in pertussis testing and surveillance, including the widespread introduction of multiplex PCR testing, alongside the larger surge in cases in England following the COVID-19 pandemic.

## Data Availability

The data that support the findings of this study have been assessed by the UK Health Security Agency Office for Data Acquisition and Release as having sensitive personal information, whose publication may breach the ISB1523: Anonymisation standard for publishing health and social care data. Therefore, to protect the privacy of participants, we cannot make this data publicly available. However, some summaries of the data may be available upon reasonable request from the UKHSA. Requests can be directed to.
The analysis presented in this paper was based on enhanced, national surveillance data. Ethics committee approval, and individual patient consent, for the work was not required because UKHSA (the organization to which all authors on the paper are affiliated) has permission to collect and process confidential patient information under regulation 3 of the Health Service (Control of Patient Information) Regulations 2002 in the UK (www.legislation.gov.uk/uksi/2002/1438/regulation/3/made).

## Declarations

## Acknowledgements

We thank UKHSA Health Protection Teams and clinical staff working in the NHS who have supported the national surveillance, as well as UKHSA and NHS laboratories for providing information to support this analysis.

## Data availability

The data that support the findings of this study have been assessed by the UK Health Security Agency Office for Data Acquisition and Release as having sensitive personal information, whose publication may breach the ISB1523: Anonymisation standard for publishing health and social care data. Therefore, to protect participants’ privacy, we cannot make this data publicly available. However, some summaries of the data may be available upon reasonable request from the UKHSA. Requests can be directed to.

## Ethics

Pertussis surveillance in England is undertaken under regulation 3 of the Health Service (Control of Patient Information) Regulations 2002 to collect confidential patient information (www.legislation.gov.uk/uksi/2002/1438/regulation/3/made) under sections 3(i) (a) to (c), 3(i)(d) (i), and (ii) and 3(3).

